# Estimating the risk of COVID-19 death during the course of the outbreak in Korea, February- May, 2020

**DOI:** 10.1101/2020.03.30.20048264

**Authors:** Eunha Shim, Kenji Mizumoto, Wongyeong Choi, Gerardo Chowell

## Abstract

Background: In Korea, a total of 10,840 confirmed cases of COVID-19 including 256 deaths have been recorded as of May 9, 2020. The time-delay adjusted case fatality risk (CFR) of COVID-19 in Korea is yet to be estimated. Methods: We obtained the daily series of confirmed cases and deaths in Korea reported prior to May 9, 2020. Using statistical methods, we estimated the time-delay adjusted risk for death from COVID-19 in Daegu, Gyeongsangbuk-do, other regions in Korea, as well as the entire country. Results: Our model-based crude CFR fitted the observed data well throughout the course of the epidemic except for the very early stage in Gyeongsangbuk-do; this was partially due to the reporting delay. Our estimates of the risk of death in Gyeongsangbuk-do reached 25.9% (95% CrI: 19.6%-33.6%), 20.8% (95% CrI: 18.1%-24.0%) in Daegu and 1.7% (95% CrI: 1.1%-2.5%) in other regions, whereas the national estimate was 10.2% (95% CrI: 9.0%-11.5%). Conclusions: The latest estimates of CFR of COVID-19 in Korea are considerably high, even with the early implementation of public health interventions including widespread testing, social distancing, and delayed school openings. Geographic differences in the CFR are likely influenced by clusters tied to hospitals and nursing homes.

## 1. Introduction

Since the first reports of cases from Wuhan in the Hubei Province of China in December 2019, more than 84,400 cases of the 2019 novel coronavirus disease (COVID-19), including 4,643 deaths have been reported in China [1]. Based on the estimates by a joint fact-finding mission of the World Health Organization (WHO) and Chinese authorities, the epidemic in China peaked between late January and early February 2020 [2]. However, the virus has been seeded in numerous countries and is now causing sustained outbreaks in several areas, with the rate of new cases outpacing that in China. As of May 9, 2020, the global cumulative number of reported infections and deaths were 3,759,967 and 259,474, respectively [1]. Outside China, new cases initially occurred primarily among travelers from China and their contacts, but local transmission has driven major outbreaks in numerous countries including the U.S., Spain, Russia, and U.K. [1].

The onset of the COVID-19 outbreak in South Korea was on January 19, 2020, when the first confirmed infected subject entered the country from Wuhan, China [3]. The severe acute respiratory syndrome coronavirus-2 (SARS-CoV-2) has since continued to spread in Korea in the form of a series of clusters of variable size and geographic location in addition to imported cases [4]. As of May 9, 2020, the cumulative number of COVID-19 reported cases and deaths in Korea was 10,840 and 256, respectively [5].

To accurately estimate the epidemiological and economic burden of an infectious disease, assessing the associated risk of death as well as the transmission potential are critical. Although the transmission modes of COVID-19 are not completely understood, person-to-person spread through respiratory droplets appears to be the main mode of transmission. The transmissibility of COVID-19 was found to be relatively high, with the mean basic reproduction number in the range 1.4-1.6 in Korea [4] and 2-7.1 in China [6-13]. In the absence of vaccines against COVID-19 or antiviral drugs for its treatment, epidemic control relies on the implementation of symptom monitoring and non-pharmaceutical interventions such as social distancing and the use of face masks.

Assessing the severity of infection of COVID-19 facilitates the prediction of the risk of death during the course of the epidemic. The crude case fatality risk (CFR) can be measured by estimating the proportion of the cumulative number of deaths to the cumulative number of cases at a certain point in time. However, owing to the delay from diagnosis to death, this metric cannot accurately capture the increase in the number of fatal cases and therefore underestimate infection severity. On the other hand, the denominator is calculated based on confirmed cases only, and thus the actual CFR based on all infected individuals may be overestimated, resulting in ascertainment bias.

As Korea has instituted large-scale testing at the core of the control interventions [14], the aim of the present study is to estimate the risk of death among confirmed cases, considering the ascertainment bias and right-censored likelihood for modeling the count of deaths by using established methods [15]. Specifically, to estimate the current severity of the COVID-19 epidemic in Korea, we report the estimates of the time-delay adjusted CFR for Daegu, Gyeongsangbuk-do, other regions (i.e., outside Daegu and Gyeongsangbuk-do), and Korea (national), with quantified uncertainty.

## 2. Method

### 2.1 Data sources

We obtained the daily series of confirmed cases and deaths in Korea from daily reports published by the Korea Centers for Disease Control and Prevention (KCDC) [5]. These data were categorized by geographic area, namely, Daegu, Gyeongsangbuk-do, other regions, and Korea (national), considering that 70% of the deaths are currently occurring in Daegu. The virus that causes COVID-19, that is, SARS-CoV-2 RNA, can be detected by reverse-transcription polymerase chain reaction (RT-PCR), and according to the Center for Laboratory Control of Infectious Diseases in KCDC, the upper limit of the Ct value for positive RT-PCR of SARS-CoV-2 is 35, whereas the negative criterion is a Ct value of 37 or higher [16]. Our analysis relies on epidemiological data reported prior to May 9, 2020.

### 2.2 Case fatality ratio

The crude CFR is defined as the number of cumulative deaths divided by the number of cumulative cases at a specific point in time. For the real-time estimation of CFR, we employ the delay *h_s_* from hospitalization to death. This is assumed to be given by *h_s_* = *H(s) – H*(*s-*1) for *s* > 0 where *H*(*s*) is the cumulative density function of the delay from hospitalization to death and follows a gamma distribution with a mean of 10.1 days and SD of 5.4 days, according to a recent study that estimated the CFR in China [17]. We let *π_a,ti_* be the time-delay adjusted CFR for reported day *t_i_* in area *a*. The likelihood function of the estimate *π_a,ti_* is calculated as

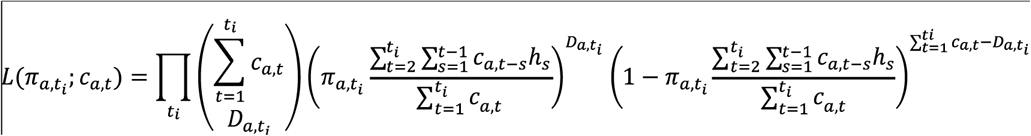

where *c_a,t_* represents the number of new cases reported on day *t* in area *a*, and *D_a,ti_* is the cumulative number of deaths until day *t_i_* in area *a* [17-19]. Among the cumulative cases with reported day *t* in area *a, D_a,ti_* have died, and the remainder have survived the infection. The factor in the second parenthesis represents the contribution of those who have died (with biased death risk), and the factor in the third parenthesis represents the contribution of the survivors. We assume that *D_a,ti_* is the result of a binomial sampling process with probability *π_a,ti_*.

We estimated the model parameters using the Monte Carlo Markov Chain (MCMC) method in a Bayesian framework. The posterior distributions of the model parameters were estimated by sampling from the three Markov chains. For each chain, we drew 100,000 samples from the posterior distribution after a burn-in of 20,000 iterations. The convergence of the MCMCs was evaluated using the potential scale reduction statistic [20,21]. The estimates and 95% credibility intervals are based on the posterior probability distribution of each parameter and on the samples drawn from these distributions. The statistical analysis was conducted in R version 3.6.1 (R Foundation for Statistical Computing, Vienna, Austria) using the rstan package.

## 3. Results

As of May 9, 2020, a total of 10,840 COVID-19 cases and 256 deaths have been reported in Korea. Among these, 6,859 cases (63.3 %) are from Daegu, and 1,366 cases (12.6 %) are from Gyeongsangbuk-do (Table 1). Among the 256 deaths, 178 (69.5%) are from Daegu, 53 (20.7%) are from Gyeongsangbuk-do and only 25 (9.8%) are from other areas. It is noted that the fatality risk increases dramatically with age, and the oldest age group exhibits the highest case fatality (Table 2). As in other affected countries including China and the U.S., it has been reported that males have higher mortality than females [22,23].

**Table 1.** Time-delay adjusted CFR of COVID-19 in the three areas under study (as of May 9th, 2020)

| Area | Latest estimate | Range of median estimates during the study period | Crude CFR |
| --- | --- | --- | --- |
| Daegu | 20.8% (95% CrIs: 18.1%-24.0%) | 1.2%-20.8% | 2.6% (95% CI<sup>‡</sup>: 2.2%-3.0%)<br>178/6859 |
| Gyeongsangbuk-do | 25.9% (95% CrI: 19.6%-33.6%) | 2.0%-83.6% | 3.9% (95% CI: 2.9%-5.0%)<br>53/1366 |
| Other regions | 1.7% (95% CrI: 1.1%-2.5%) | 0.6%-3.8% | 1.0% (95% CI: 0.6%-1.4%)<br>25/2615 |
| Korea (national) | 10.2% (95% CrI: 9.0%-11.5%) | 1.3%-16.6% | 2.4% (95% CI: 2.1%-2.7%)<br>256/10840 |

<sup>§</sup>CrI: 95% credibility intervals (CrI), <sup>‡</sup>95%CI: 95% confidence interval

**Table 2.** Distribution of the cases by sex and age group (as of May 9, 2020) [5]

|  |  | Confirmed cases # (%) | Deaths # (%) | Fatality rate (%) |
| --- | --- | --- | --- | --- |
| **Total** |  | **10,840 (100.00)** | **256 (100.00)** | **2.36** |
| Sex | Male | 4,406 (40.65) | 133 (51.95) | 3.02 |
|  | Female | 6,434 (59.35) | 123 (48.05) | 1.91 |
| Age Group | 0-9 | 141 (1.30) | 0 (0.00) | - |
|  | 10-19 | 594 (5.48) | 0 (0.00) | - |
|  | 20-29 | 2,979 (27.48) | 0 (0.00) | - |
|  | 30-39 | 1,177 (10.86) | 2 (0.78) | 0.17 |
|  | 40-49 | 1,438 (13.27) | 3 (1.17) | 0.21 |
|  | 50-59 | 1,958 (18.06) | 15 (5.86) | 0.77 |
|  | 60-69 | 1,355 (12.50) | 37 (14.45) | 2.73 |
|  | 70-79 | 710 (6.55) | 77 (30.08) | 10.85 |
|  | 80 and above | 488 (4.50) | 122 (47.66) | 25.00 |

The cumulative cases in (A) Korea (national), (B) Daegu, (C) Gyeongsangbuk-do, and (D) other regions, and the cumulative deaths in (E) Korea (national), (F) Daegu, (G) Gyeongsangbuk-do and (H) other regions are shown in Figure 1. The curve of the cumulative number of deaths grows after that of the cumulative number of cases, and the increase in the number of deaths in Daegu is more rapid; furthermore, the associated mortality burden appears to be significantly higher than in the rest of Korea.

**Figure 1.**
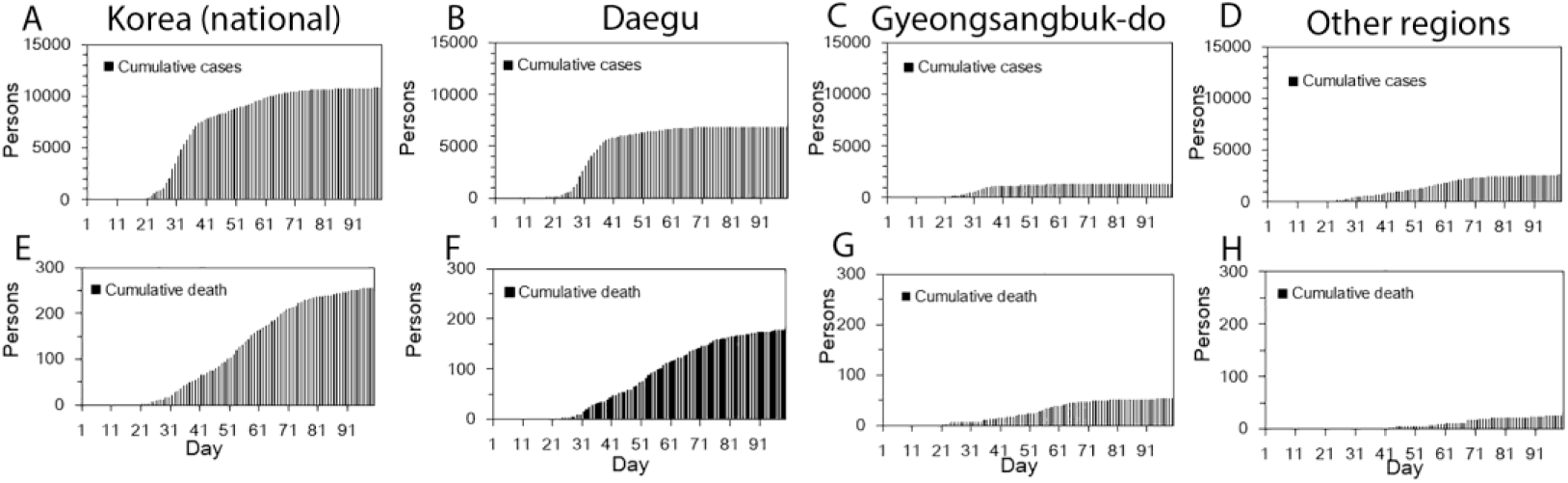
Temporal distribution of cases and deaths by area, February – May 2020. Cumulative cases in (A) Korea (total), (B) Daegu, (C) Gyeongsangbuk-do, and (D) other regions, and cumulative deaths in (E) Korea (total), (F) Daegu, (G) Gyeongsangbuk-do, and (H) other regions. Day 1 corresponds to February 1^st^, 2020. As the dates of illness onset were not available, the dates of reporting were used.

The observed and model-based posterior estimates of the crude CFR in (A) Korea (national), (B) Daegu, (C) Gyeongsangbuk-do, and (D) other regions, and the model-based posterior estimates of the time-delay adjusted CFR in (E) Korea (national), (F) Daegu, (G) Gyeongsangbuk-do, and (H) other regions, for February – May, 2020 are shown in Figure 2. Our model-based crude CFR fitted the observed data well throughout the course of the epidemic except for the very early stage in Gyeongsangbuk-do, probably owing to the reporting delay of confirmed cases. During the course of the outbreak, our model-based posterior estimates of the time-delay adjusted CFR take considerably higher values than the observed crude CFR. Our estimates of the time-delay adjusted CFR appear to be decreasing at the early stage almost consistently in all three areas, whereas in Gyeongsangbuk-do, the estimates were low at the early stage, subsequently increased, peaked in the midst of the study period, and are now following an increasing trend similar to those in the other two areas.

**Figure 2.**
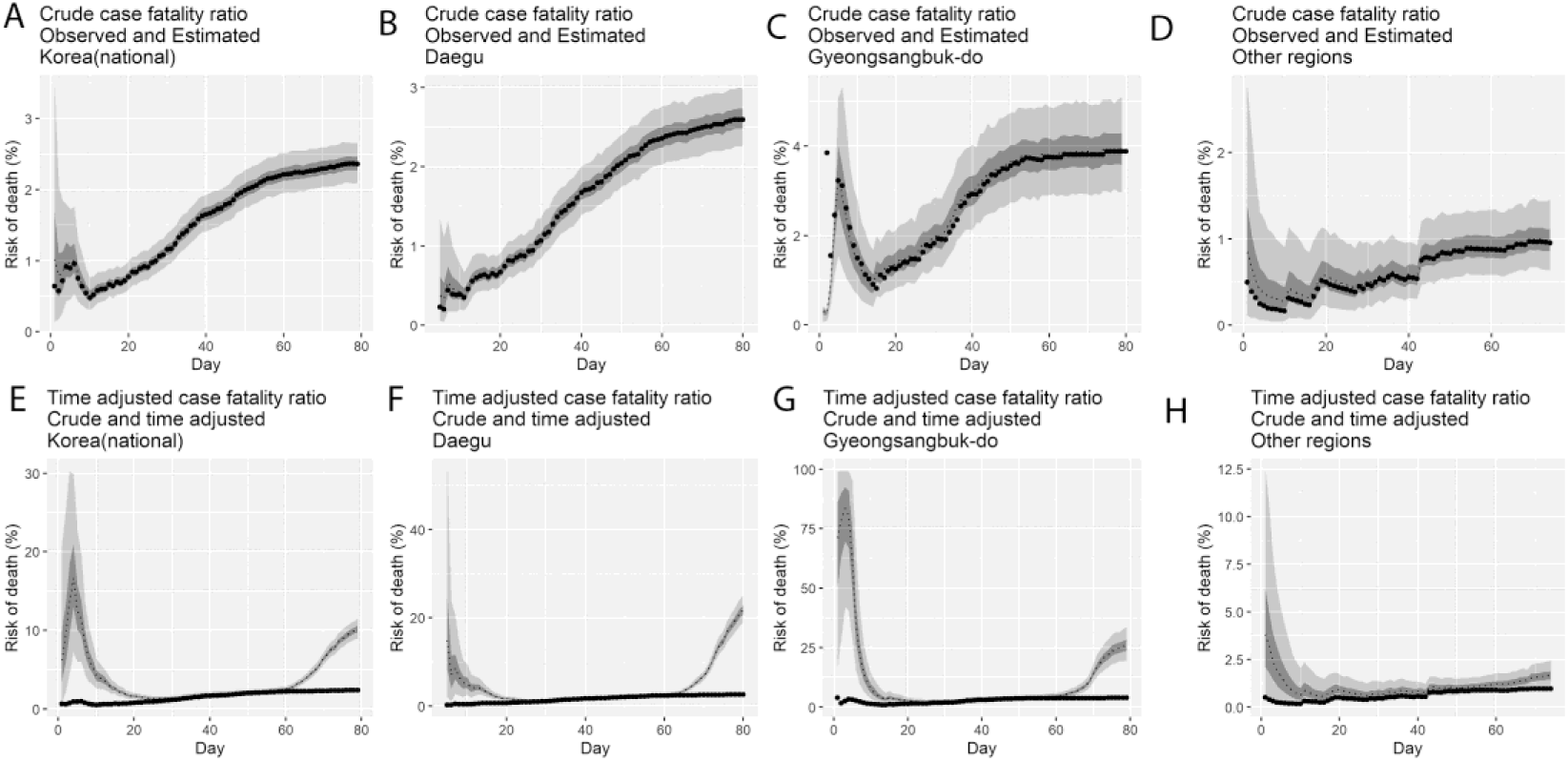
Temporal variation of risk of death, Korea, February – May 2020. Observed and posterior estimates of the crude CFR in (A) Korea (national), (B) Daegu, (C) Gyeongsangbuk-do, and (D) other regions, and time-delay adjusted CFR in (E) Korea (national), (F) Daegu, (G) Gyeongsangbuk-do, and (H) other regions. Day 1 corresponds to February 1^st^, 2020. Black dots represent the crude CFR, the light and dark colors indicate 95% and 50% credible intervals for the posterior estimates, respectively.

The latest estimates (May 9, 2020) of the time-delay adjusted CFR are 20.8% (95% CrI: 18.1%-24.0%) in Daegu, 25.9% (95% CrI: 19.6%-33.6%) in Gyeongsangbuk-do, 1.7% (95% CrI: 1.1%-2.5%) in other regions and 10.2% (95% CrI: 9.0%-11.5%) in Korea (national), whereas the observed crude CFR is 2.6% (95% CI: 2.2%-3.0%) in Daegu, 3.9% (95% CI: 2.9%-5.0%) in Gyeongsangbuk-do, 1.0% (95% CrI: 0.6%-1.4%) in other regions, and 2.4% (95% CrI: 2.1%-2.7%) in Korea (national) (Figure 3, Table 1).

**Figure 3.**
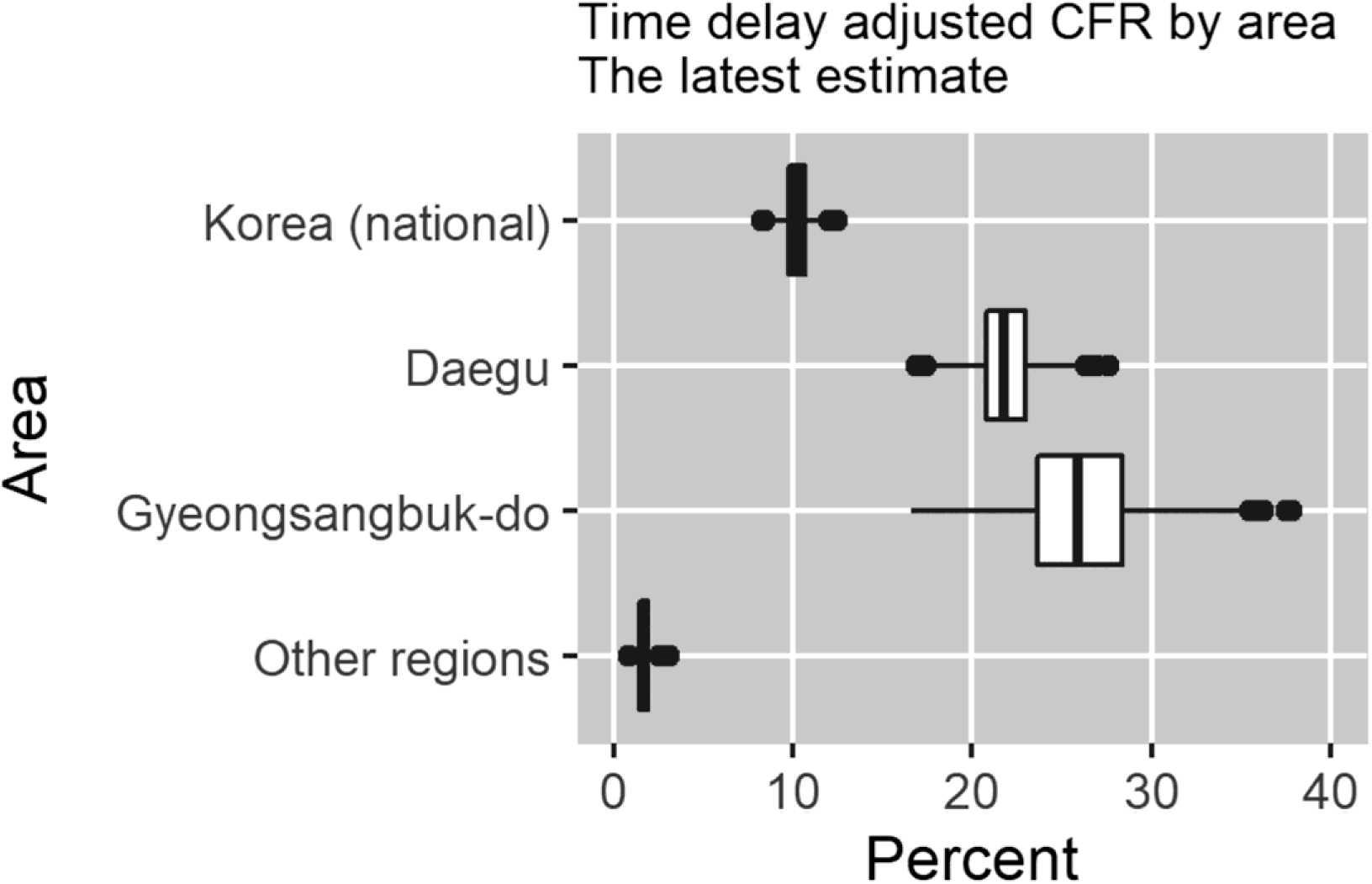
Latest estimates of time-delay adjusted CFR of COVID-19 by area (May 9, 2020).

## 4. Discussion

In this study, we derived estimates of the CFR for the ongoing COVID-19 epidemic in Korea, which is heavily relying on intensive social distancing and mass testing of suspected COVID-19 cases to mitigate the incidence levels of the disease and deliver early medical care to the most vulnerable so that the risk of death may be reduced. Specifically, we estimated the time-delay adjusted CFR in three different regions, and we demonstrated that the most severely affected areas were Gyeongsangbuk-do and Daegu, whereas the rest of Korea exhibited a less severe profile. The delay-adjusted CFR in Gyeongsangbuk-do, largely driven by hospital-based transmission, was considerably high, estimated at 25.9% (95% CrI: 19.6%-33.6%), which was approximately 2.5-fold greater than our estimate for all regions in Korea.

The fatality distribution indicates the age-dependency of COVID-19. Accordingly, it is highly important to monitor infected individuals of advanced age and/or underlying medical conditions. It has also been reported that the risk of symptomatic infection increases with age, although this may be partially affected by the preferential ascertainment of older and thus more severe cases [24]. In fact, a substantial fraction of the case patients were elderly or had underlying conditions [25]. The youngest case patient was a 35-year old Mongolian male who already had complications from chronic hepatic failure with cirrhosis and esophageal variceal bleeding [25]. The second-youngest case patient was a 39-year old female with cerebral hemorrhage due to arteriovenous malformation, and she was diagnosed with COVID-19 on March 3, 2020 [26]. Except for these two cases, all deaths associated with COVID-19 in Korea occurred among individuals of age over 40.

Our estimated time-adjusted CFR associated with COVID-19 in all regions in Korea is 10.2% (95% CI: 9.0%-11.5%). This estimate is higher than the reported CFR for other coronaviruses such as SARS-CoV [27] and the Middle East respiratory syndrome (MERS) coronavirus [28], as well as the estimates from the 2009 H1N1 influenza pandemic [29,30]. In addition, our estimates are relatively higher than the CFR obtained from affected areas in China (ranging from 1.41% to 5.25%), based on the cases and deaths observed to date [17,31]. In general, the CFR of the same disease may vary greatly in different countries or even different regions of the same country, partially owing to differences in health control policies, availability of healthcare, medical standards, and detection efficiency. In Korea, an extensive COVID-19 testing regime has been instituted, and a massive contact tracing was performed as case numbers grew. As a result, the country has tested more people per capita than any country in the world, with a total of 660,030 people as of May 9, 2020 [5]. Even with such extensive testing that may have effectively identified COVID-19 cases, the CFR was shown to be considerably high in Korea.

The reported CFR of COVID-19 tends to vary over the course of the epidemic. It is noteworthy that the upward trend of the CFR during the early phase generally indicates increasing ascertainment bias. During a growing epidemic, the final clinical outcome of most reported cases is unknown, and thus the true CFR is underestimated early in the epidemic. This was observed in our results (Figure 2) as well as in prior studies of epidemics of respiratory pathogens including SARS and H1N1 influenza [32,33]. Similarly, the observed time lag between symptom onset and clinical outcomes tends to bias the estimated CFR downward during the early growth phase of an epidemic.

When the transmission of COVID-19 was widely recognized by clusters of confirmed cases such as hospital-based transmission, surveillance was typically biased toward detecting clinically severe cases, therefore resulting in higher CFR estimates, as shown in Figure 2 as the epidemic progressed. Specifically, the virus has generated COVID-19 outbreaks in hospitals and nursing homes, including clusters based in Cheongdo Daenam Hospital (resulting in 120 infections), Bonghwa Pureun Nursing Home (68 infections), and Gyeongsan Seo Convalescent Hospital (66 infections) in Gyeongsangbuk-do; this explains our higher CFR estimate at 25.9% (95% CrI: 19.6-33.6%) for Gyeongsangbuk-do. Similarly, nosocomial clusters also occurred in Uijeongbu St. Mary’s Hospital (50 infections) in Gyeonggi province, Second Mi-Ju Hospital (196 infections), Hansarang Convalescent Hospital (128 infections), and Daesil Convalescent Hospital (100 infections) in Daegu [5].

Such hospital-based transmission resulted in secondary infections affecting healthcare workers, as well as inpatients and their visitors, thus elevating CFR estimates, as has been previously documented for outbreaks of MERS and SARS in the past [34,35]. In addition, a large number of COVID-19 cases are related to church clusters in Korea, including a total of 5,212 COVID-19 cases in a cluster linked to the Shincheonji Church, accounting for approximately 48% of all confirmed cases [5].

Asymptomatic patients complicate the estimation of the CFR for COVID-19. In fact, the transmission of SARS-CoV-2 from asymptomatic individuals (or individuals in the incubation period) has also been described [36-38]. For instance, in a COVID-19 outbreak on a cruise ship, approximately half of the 619 confirmed cases were asymptomatic at the time of diagnosis [39]. Moreover, some patients with asymptomatic infection exhibited objective clinical abnormalities. In a previous study of 24 patients with asymptomatic infection, chest computed tomography indicated that 50% of the patients had typical ground-glass opacities or patchy shadowing, and another 20% had atypical imaging abnormalities [40]. In addition, it is likely that asymptomatic or very mild cases were not reported, and thus they were not included in our analysis, possibly overestimating the CFR. Future research and more accurate incidence data including age-stratified serologic studies would improve our estimates.

## Data Availability

Data are publicly available.

https://www.cdc.go.kr/board/board.es?mid=a30402000000&bid=0030

## Author Contributions

ES, KM and GC analyzed the data. WC retrieved and managed the data. ES, KM, and GC wrote the first draft of the manuscript. All authors contributed to the writing of the manuscript, and have read and agreed to the published version of the manuscript.

## Funding

This work was supported by the National Research Foundation of Korea (NRF) grant funded by the Korea government (MSIT) [No. 2018R1C1B6001723] to ES and WC. KM acknowledges support from the Japan Society for the Promotion of Science (JSPS) KAKENHI Grant Number 15K20936, from Program for Advancing Strategic International Networks to Accelerate the Circulation of Talented Researchers Grant Number G2801 and from the Leading Initiative for Excellent Young Researchers from the Ministry of Education, Culture, Sport, Science & Technology of Japan. GC acknowledges support from NSF grant 1414374 as part of the joint NSF-NIH-USDA Ecology and Evolution of Infectious Diseases program, and UK Biotechnology and Biological Sciences Research Council grant [BB/M008894/1].

## Acknowledgments

None.

## Conflicts of Interest

We declare no competing interests.

## Notes

### Competing Interest Statement

The authors have declared no competing interest.

